# Factors Associated with Recurrent Malaria Episodes among Children Under Five at Kayunga Regional Referral Hospital in Kayunga District, Central Uganda

**DOI:** 10.1101/2025.02.15.25322352

**Authors:** Derick Modi, Marvin Musinguzi, Patricia Pita, Eustes Kigongo, Amir Kabunga, Julius Kayizzi, Deo Kasaija, Voni Alice Khanakwa, Oscar Simon Alyao, Julius Lubangakene, Tom Murungi, Christopher Okullo Oneka, Marc Sam Opollo

## Abstract

**Background:** Malaria poses a substantial global challenge and continues to be a major cause of mortality and morbidity in numerous developing nations. Children under the age of five in low- and middle-income countries such as Uganda are the most affected. Recurrent episodes of malaria have significant consequences for both children and their families. However, there remains a deficiency in knowledge regarding recurrent malaria episodes in Uganda.

**Objective:** To determine the prevalence and factors associated with recurrent malaria episodes among children under five at Kayunga Regional Referral Hospital.

**Methodology:** This was a cross-sectional study conducted among children under five at Kayunga Regional Referral Hospital in central Uganda. The data was collected among 254 consecutively sampled participants who were caring for children under five. Data was collected using a researcher-administered questionnaire and analyzed at univariate, bivariate, and multivariate levels.

**Results:** A total of 250 participants participated in the study with a response rate of 98.45%. The prevalence of recurrent malaria episodes was 84% (210). The factors associated with recurrent malaria episodes were; children from houses that were annually sprayed (aOR; 8.93, 95CI%,2.11-37.81), children who were treated with quinine antimalarial in the previous infection (aOR, 0.28, 95%CI,0.12-0.65) and children who were residing in a house whose windows were closed at 7-8 pm (aOR, 8.31, 95%CI, 2.21-31.27).

**Conclusion:** Nearly 90% of children under the age of five years experienced recurrent malaria episodes. These recurrent infections were more frequent among children from houses that were annually sprayed and those from houses with delayed window closure compared to those who had been treated using quinine-based antimalarials. While quinine-based antimalarials remain an important treatment option, alternative or complementary malaria prevention strategies, such as frequent indoor spraying and early closure of windows should be prioritized.

## Background

Malaria remains one of the leading public health burdens in the world despite the remarkable achievements made towards its control and prevention since the beginning of the second millennium [1]. Globally, according to the World Malaria Report 2020, there were an estimated 241 million cases of malaria and 627,000 malaria. In Africa, four African countries accounted for just over half of all malaria deaths worldwide: Nigeria (31.9%), the Democratic Republic of the Congo (13.2%), the United Republic of Tanzania (4.1%), and Mozambique at 3.8% [2]. Uganda has the 3^rd^ highest proportion of malaria cases (5%) and the 8^th^ highest level of deaths (3%)[3]. It also has the highest proportion of malaria cases in East and Southern Africa, 23.7% among children under five [4]. According to the WHO children under the age of 5 years are the most affected reporting that more than two-thirds of deaths are due to malaria [5]. The recurrent malaria episodes in children under five present a higher risk of mortality because children under five have an underdeveloped immune system compared to adults. This makes them less capable of fighting off infections like malaria. Recurrent infections can further weaken their immune responses, leading to higher vulnerability and risk of severe complications [6].

A recurrent malaria infection is a newly detectable episode of blood-stage parasitemia occurring after a previous infection [7]. This can be due to a relapse of *P. vivax* and *P. ovale* infections [2] and usually occurs after 48 to 72 hours when the parasites come out of hibernation and begin invading red blood cells [7]. The prevalence of recurrent malaria infection can vary depending on various factors such as the geographical location, intensity of malaria transmission, access to healthcare, and the effectiveness of malaria control interventions [8]. Recurrent episodes of malaria among children under five can contribute to poor nutrition, weight loss, and stunted growth [9]. Malnutrition diminishes a child’s immune system, rendering them more vulnerable to subsequent infections[10]. On the other hand, Emotional and psychological consequences include Chronic illness, such as recurrent episodes of malaria, which can have a negative impact on a child’s emotional well-being. [11].

Uganda has the sixth highest number of annual deaths from malaria in Africa, as well as some of the highest reported malaria transmission rates in the world, with approximately 16 million cases reported in 2013 and over 10,500 deaths annually. In addition, malaria has an indirect impact on the economy and development in general [12]. However, clinically diagnosed malaria is the leading cause of morbidity and mortality, accounting for 30-50% of outpatient visits at health facilities, 15-20% of all hospital admissions, and up to 20% of all hospital deaths [13]. The government of Uganda has made efforts to provide LLITN to households and IRS to control malaria infections in children under five [14]

Malaria remains a significant public health challenge, particularly among children under five in sub-Saharan Africa. Despite extensive research and interventions, recurrent malaria episodes persist, exacerbating child morbidity and mortality. Kayunga has a malaria prevalence of 18.4% according to the Malaria Indicator Survey 2020 [15]. At Kayunga Regional Referral Hospital, the factors contributing to these recurrent episodes are not well understood, highlighting a critical gap in the literature. Existing studies primarily focus on initial malaria incidence and general prevention measures [16–18], with limited attention to the specific determinants of recurrence in this vulnerable age group. Factors such as socio-economic conditions, healthcare access, and specific local epidemiological characteristics have not been comprehensively examined in the context of Kayunga. Addressing these gaps is crucial for developing targeted strategies to reduce the burden of recurrent malaria and improve health outcomes for children under five in this region. We thus determined the prevalence and factors associated with recurrent malaria episodes among children under five at Kayunga Regional Referral Hospital, Kayunga district in central Uganda.

## Materials and methods

### Study design and setting

We employed a cross-sectional study design that used quantitative methods to collect and analyze the data. The study was conducted from 10^th^-25^th^ March 2024. The study was conducted at Kayunga Regional Referral Hospital’s Outpatient Department and the paediatric ward. Kayunga district is located in Central Uganda and has a grid reference of 0° 59’ 9.6648’’ N and 32° 51’ 12.8736’’ E. It has an elevation of 1063m(3488ft). Kayunga District is 74 Km East of Kampala City, bordered by Mukono district to the south, Jinja and Buikwe to the east, Kamuli to the northeast, Amolator & Apac in the North, Luwero in the west and Nakasongola to the northwest.

Kayunga district has a malaria prevalence of 18.4%, according to the Malaria Indicator Survey 2020 [20]. However, In Kayunga, little is known about recurrent malaria episodes among children under five.

### Study population

The study population consisted of caretakers, parents, or guardians of children under five who came to access care at Kayunga Regional Referral Hospital. We recruited caretakers, parents, or guardians of children under five who had consented and were available at the paediatric ward or outpatient clinic of Kayunga Regional Referral Hospital at the time of data collection. We excluded caretakers who had not spent at least one month with the children before data collection.

### Sample size and sampling technique

The sample size was determined using the Kish Leslie (1965) formula (n=Z^2^PQ/d^2^) [19]. Z=1.96%, d=0.05, p=18.4%, which is the malaria prevalence among children under five according to the Uganda malaria indicator survey. This resulted in a sample size of 231. When adjusted for a non-response rate of 10%, an overall sample size of 254 participants was generated.

A consecutive sampling method was used to select participants for data collection. The study included all caregivers of children under the age of five who met the inclusion criteria and sought health care at Kayunga Regional Referral Hospital’s Outpatient Department (OPD) and the paediatric ward.

### Data collection tools

Data was collected using a researcher-administered questionnaire. The questionnaire was adapted from Simões and colleagues and tailored to the study setting [20]. Questions were divided into three sections, including socio-demographics (age, religion, economic status, etc.), household factors (Poor housing construction, lack of mosquito control, poor sanitation and hygiene, stagnant water, lack of knowledge and awareness), and practices (use of insecticide-treated bed nets, indoor residual spraying, larval source management, self-medication, over using antimalarial). Before the actual data collection, the tool was also pretested at Lira University Teaching Hospital and translated to Luganda, the local language spoken by the majority of residents at Kayunga Regional Referral Hospital.

We defined recurrent malaria episodes as having suffered from at least two or more episodes of malaria infection within 30 days, with at least one episode being diagnosed using a rapid diagnostic test (RDT) or microscopy [21]. Thus, it was measured using a binary response (yes/no), where those whose children suffered from at least two or more episodes of malaria infection within 30 days, with at least one episode being diagnosed using a rapid diagnostic test (RDT) or microscopy were categorised as “yes”. On the other hand, those whose children had not suffered at least two malaria episodes or those whose malaria was not medically tested and diagnosed were categorised as “no”.

### Data collection procedure

Upon ethical clearance from the Lira University Research Ethics Committee, the researcher sought permission from the District Health Officer, the principal medical officer, and other relevant authorities at Kayunga Regional Referral Hospital to conduct the study. The researcher then trained five research assistants to collect data for the participants who met the eligibility criteria, and data collection was done by conducting researcher-administered interviews using semi-structured questionnaires after getting consent from the participants. Before the data collection process, written informed consent to participate in the study was sought from the participants. Recurrent malaria infection was self-reported by the caretakers of the children and was validated by asking the caretakers to provide at least one record of a previous malaria diagnosis from a hospital or clinic.

### Data management and analysis

Data was entered, cleaned and analysed in SPSS version 26. During univariate analysis, descriptive statistics were used to summarise the data and determine the prevalence of recurrent malaria episodes including frequencies and percentages. At bivariate analysis, a bivariate logistic regression was performed between the independent variables and dependent variable at a 95% confidence interval, Crude odds ratios (COR) were used as measures of association. Variables with P ≤0.05 were considered as having significant associations with the dependent variable. In multivariate analysis, variables with P ≤0.2 at the bivariate level were included in the multivariate logistic regression at a 95% confidence interval. Adjusted Odds Ratios were used to measure association. Variables that had P ≤0.05 at multivariate logistic regression were considered to be significantly associated with the dependent variable.

### Ethical considerations

Ethical approval to conduct the study was obtained from the Lira University Research Ethics Committee and approval was issued under rec number LUREC-2023-66. Upon ethical clearance from the Lira University Research Ethics Committee.

Administrative clearance to conduct the study was sought from the principal medical officer, and other relevant authorities at Kayunga Regional Referral Hospital. Written informed consent was sought from all the participants, in this study, and these were mothers, and guardians of the children after sharing with them the objectives of the study, possible benefits and risks of the study. Identifiers such as names, phone numbers, or personal addresses were not included in the questionnaires to protect the participants’ privacy. Codes were used to identify participants. The hard copy of the questionnaires was kept under lock and key and only accessible by the research team.

## Results

Out of the 254 participants in the study, data was only collected from 250 participants at Kayunga Regional Referral Hospital in Central Uganda. This resulted in a response rate of 98.45%.

### Recurrent malaria episodes and socio-demographic characteristics of the respondents

Among the socio-demographic variables, the following were significantly associated with recurrent malaria episodes. Children who were not in school experienced many malaria episodes (cOR;0.50,95%CI,0.252-0.976, P=0.04), children whose parents had a monthly income of 330,000-930,000ugx, were more likely to experience recurrent malaria episodes (cOR;2.83,95%CI,1.098-7.287,p=0.03), children who had underlying conditions had many recurrent episodes of malaria (cOR;2.69,95%CI,1.30-5.55,p=0.01), children who had other underlying conditions had more malaria episodes(cOR;2.31,95%CI,0.50-10.65, P=0.01).

**Table 1:** Showing the Bivariate analysis results of the socio demographic characteristics associated with recurrent malaria episodes among children under five at Kayunga Regional Referral Hospital.

| Variable | Frequency<br>N (%) | Malaria reoccurrence |  | cOR (95% CI) | P value. |
| --- | --- | --- | --- | --- | --- |
|  |  | **No**<br>**reoccurrence** | **Reoccurrence** |  |  |
| **Malaria recurrent episodes** |  | **84%** | **16%** |  |  |
| **Age of the participant.** |  |  |  |  |  |
| 15-34 | 3 (1.20) | 1(33.30) | 2(66.70) | Ref |  |
| 35-54 | 128(51.20) | 21(16.40) | 107(83.60) | 0.57(0.41-8.05) | 0.681 |
| 55-74 | 101(40.40) | 15(14.90) | 86(85.10) | 1.46(0.44-4.86) | 0.542 |
| 75-above | 18(7.20) | 4(22.20) | 14(77.80) | 1.64(0.47-5.66) | 0.440 |
| **Age of the child in months.** |  |  |  |  |  |
| 3-12, infant | 61(24.40) | 12(19.70) | 49(80.30) | Ref |  |
| 13-48, toddler | 162(64.80) | 24(14.80) | 138(85.20) | 0.93(0.29-2.96) | 0.900 |
| 49-above, children. | 27(10.80) | 5(18.50) | 22(81.50) | 1.31(0.45-3.79) | 0.621 |
| **Gender** |  |  |  |  |  |
| Male | 115(46.00) | 20(17.40) | 95(82.60) | Ref |  |
| Female | 135(54.00) | 21(15.60) | 114(84.40) | 0.88(0.45-1.71) | 0.701 |
| **Marital status** |  |  |  |  |  |
| Married | 132(52.80) | 20(15.20) | 112(84.80) | Ref |  |
| Cohabiting | 49(19.60) | 8(16.30) | 41(83.70) | 1.30(0.60-2.80) | 0.501 |
| Single | 69(27.60) | 13(18.80) | 56(81.20) | 1.19(0.45-3.13) | 0.732 |
| **Religion** |  |  |  |  |  |
| Catholics | 55(22.00) | 10(18.20) | 45(81.80) | Ref |  |
| Protestant | 91(36.40) | 19(20.90) | 72(79.10) | 0.49(0.14-1.68) | 0.251 |
| Muslim | 63(25.20) | 8(12.70) | 55(87.30) | 0.41(0.13-1.29) | 0.132 |
| Others | 41(16.40) | 4(9.80) | 37(90.20) | 0.74(0.21-2.65) | 0.651 |
| **Education level** |  |  |  |  |  |
| Primary | 83(33.20) | 14(16.90) | 69(83.10) | Ref |  |
| Secondary | 84(33.60) | 16(19.00) | 68(81.00) | 0.67(0.22-1.99) | 0.472 |
| Tertiary | 41(16.40) | 6(14.60) | 35(85.40) | 0.574(0.20-1.69) | 0.321 |
| None | 42(16.80) | 5(11.90) | 37(88.10) | 0.79(0.22-2.82) | 0.710 |
| **Schooling status** |  |  |  |  |  |
| In school | 87(34.8) | 20(23.0) | 67(77.0) | Ref |  |
| Not in school | 163(65.2) | 21(12.9) | 142(87.1) | 0.495(0.252-0.976) | **0.040\*** |
| **Monthly household income, (\$)** | | | | | |
| 20-84 | 158(63.20) | 18(11.40) | 140(88.60) | Ref |  |
| 85-244 | 62(24.80) | 15(24.20) | 47(75.80) | 2.83(1.10-7.29) | **0.030\*** |
| 245-above | 30(12.00) | 8(26.70) | 22(73.30) | 1.14(0.42-3.09) | 0.801 |
| **Underlying condition?** |  |  |  |  |  |
| Yes | 122(48.80) | 12(9.80) | 110(90.20) | Ref |  |
| No | 128(51.20) | 29(22.70) | 99(77.30) | 2.69(1.30-5.55) | **0.010\*** |
| **Type of Underlying condition** |  |  |  |  |  |
| Sickle cell | 12(4.80) | 3(25.00) | 9(75.00) | Ref |  |
| Measles | 24(9.60) | 4(16.70) | 20(83.30) | 0.87(0.22-3.42) | 0.871 |
| Chickenpox | 18(7.20) | 2(11.10) | 16(88.90) | 1.44(0.46-4.57) | 0.531 |
| Others | 71(28.40) | 4(5.60) | 67(94.40) | 2.31(0.50-10.65) | **0.010\*** |
| No condition. | 125(50.00) | 28(22.40) | 97(77.60) | 4.84(1.62-14.42) | 0.281 |
| **Place of residence** |  |  |  |  |  |
| Rural | 163(65.20) | 24(14.70) | 139(85.30) | Ref |  |
| Urban | 87(34.80) | 17(19.50) | 70(80.50) | 1.41(0.71-2.79) | 0.331 |
| *\*level of significance at $p<0.05$, \*\*\*level of significance at $p<0.001$, $p\leq 0.05$* |  |  |  |  |  |

### Practices towards prevention of malaria

At bivariate analysis, children in the houses which were spayed annually were less likely to experience recurrent malaria episodes (cOR;0.23,95%CI,0.09-0.58, P=0.00), children who were practicing self-medication were more likely to experience recurrent malaria episodes (cOR2.25,95%CI,2.25-1.13, P=0.02), children who were using artesunate antimalaria drug were less likely to experience recurrent malaria episodes(cOR;0.23,95%CI,0.00-0.02,0.01), children staying in houses where windows were closed from 7-8 pm had more recurrent malaria episodes (cOR;2.19,95%CI,1.10-4.36, P=0.03).

**Table 2:** Showing the Bivariate analysis of the practices towards prevention of malaria with recurrent malaria episodes among children under five at Kayunga Regional Referral Hospital.

| Variable | Frequency,<br>N (%) | Malaria reoccurrence |  | cOR (95%CI) | P value. |
| --- | --- | --- | --- | --- | --- |
|  |  | No<br>reoccurrence | Re<br>occurrence |  |  |
| House sprayed with insecticide. |  |  |  |  |  |
| Yes | 81(32.40) | 17(21.00) | 64(79.00) | Ref |  |
| No | 169(67.60) | 24(14.20) | 145(85.80) | 0.62(0.31-1.24) | 0.180 |
| **Period sprayed** |  |  |  |  |  |
| Monthly | 24(9.60) | 10(45.50) | 12(54.50) | Ref |  |
| Annually | 50(20.00) | 4(8.00) | 46(92.00) | 0.23(0.09-0.58) | **<0.001\*\*\*** |
| Others | 7(2.80) | 3(42.90) | 4(57.10) | 1.91(0.63-5.77) | 0.260 |
| I don't spray | 169(67.60) | 24(14.20) | 145(85.80) | 0.221(0.05-1.05) | 0.060 |
| **Use of treated mosquito net** |  |  |  |  |  |
| Yes | 212(84.80) | 35(16.50) | 177(83.50) | Ref |  |
| No | 38(15.20) | 6(15.80) | 32(84.20) | 0.95(0.37-2.44) | 0.950 |
| **Self-medicate the child** |  |  |  |  |  |
| Yes | 133(15.20) | 15(11.30) | 118(88.70) | Ref |  |
| No | 117(46.80) | 26(22.20) | 91(77.80) | 2.25(2.25-1.13) | **0.020\*** |
| **Anti-malaria commonly used to treat malaria** |  |  |  |  |  |
| Coartem | 137(57.20) | 15(10.90) | 122(89.10) | Ref |  |
| Quinine | 64(25.60) | 20(31.30) | 44(68.80) | 0.86(0.18-4.04) | 0.851 |
| Artesunate | 28(11.60) | 4(14.30) | 24(85.70) | 0.23 (0.00-0.02) | **0.011\*** |
| p-alaxin | 21(4.40) | 2(9.50) | 19(90.50) | 0.63(0.10-3.82) | 0.610 |
| **Always use recommended anti-malarial** |  |  |  |  |  |
| Yes | 169(67.60) | 27(16.00) | 142(84.00) | Ref |  |
| No | 81(32.40) | 14(17.30) | 67(82.70) | 1.09(0.54-2.23) | 0.791 |
| **Use of mosquito repellants** |  |  |  |  |  |
| Yes | 73(29.20) | 15(20.50) | 58(79.50) | Ref |  |
| No | 177(70.80) | 26(14.70) | 151(85.30) | 0.66(0.33-1.35) | 0.260 |

| At what time do you close windows every day |  |  |  |  |  |
| --- | --- | --- | --- | --- | --- |
| 3-4 pm | 5(2.00) | 1(20.00) | 4(80.00) | Ref |  |
| 5-6 pm | 166(66.40) | 21(12.70) | 145(87.30) | 1.27(0.13-12.03) | 0.840 |
| 7-8 pm | 79(31.60) | 19(24.10) | 60(75.90) | 2.19(1.10-4.36) | **0.031\*** |
| Time to be indoors |  |  |  |  |  |
| 6-7 pm | 82(32.80) | 17(20.70) | 65(79.30) | Ref |  |
| 8-9 pm | 125(50.00) | 19(15.20) | 106(84.80) | 0.50(0.17-1.47) | 0.211 |
| 10-11 pm | 43(17.20) | 5(11.60) | 38(88.40) | 0.734(0.26-2.10) | 0.570 |
| *\*level of significance at $p<0.05$, \*\*\*level of significance at $p<0.001$, $p\leq 0.05$* |  |  |  |  |  |

### Household practices

At bivariate analysis, the children whose homes were near the bush were more likely to experience recurrent malaria episodes (cOR; 2.36, 95% CI,1.18-4.72, P=0.02).

**Table 3.** House hold practices associated with recurrent malaria episodes among children under five at Kayunga Regional Referral Hospital.

| Variable | Frequency<br>N (%) | Malaria reoccurrence |  | cOR(95%CI) | P value |
| --- | --- | --- | --- | --- | --- |
|  |  | No<br>reoccurrence | Re occurrence |  |  |
| Home Near a Bush |  |  |  |  |  |
| Yes | 180(72.00) | 23(12.80) | 157(87.20) | Ref |  |
| No | 70(28.00) | 18(25.70) | 52(74.30) | 2.36(1.18-4.72) | **0.020\*** |
| Household near a water body |  |  |  |  |  |
| Yes | 116(46.40) | 21(18.10) | 95(81.90) | Ref |  |
| No | 134(53.60) | 20(14.90) | 114(85.10) | 0.80(0.41-1.55) | 0.501 |
| **Household Number** |  |  |  |  |  |
| 3-8 | 178(71.20) | 31(17.40) | 147(82.60) | Ref |  |
| 9-14 | 58(23.20) | 7(12.10) | 51(87.90) | 1.29(0.34-4.91) | 0.710 |
| 15-above | 14(5.60) | 3(21.40) | 11(78.60) | 1.99(0.44-8.92) | 0.371 |
| **Total number of children in the home** |  |  |  |  |  |
| 1-3 | 53(21.20) | 8(15.10) | 45(84.90) | Ref |  |
| 4-6 | 150(60.00) | 27(17.90) | 124(82.10) | 0.84(0.27-2.64) | 0.770 |
| 7-above | 46(18.40) | 6(13.00) | 40(87.00) | 0.69(0.27-1.79) | 0.440 |
| **Availability of stagnant water in the home** |  |  |  |  |  |
| Yes | 67(26.90) | 11(16.40) | 56(83.60) | Ref |  |
| No | 183(73.20) | 30(16.40) | 153(83.60) | 0.99(0.47-2.13) | 0.991 |
| **Household experiences flooding** |  |  |  |  |  |
| Yes | 183(73.20) | 29(15.80) | 154(84.20) | Ref |  |
| No | 67(26.80) | 12(17.90) | 55(82.10) | 1.16(0.55-2.43) | 0.701 |
| *\*level of significance at $p < 0.05$, \*\*\*level of significance at $p < 0.001$, $p \leq 0.05$* |  |  |  |  |  |

### Factors associated with the prevalence of recurrent malaria episodes among children under five at Kayunga Regional Referral Hospital

At Multivariate analysis, children from houses that were annually sprayed were more likely to experience recurrent malaria episodes, (aOR; 8.93,95CI%,2.11-37.81, p= 0.003), children who were using quinine antimalarial were less likely to experience recurrent malaria episodes (aOR, 0.28,95%CI,0.12-0.65, P=0.003), children who were residing in a house whose windows were closed at 7-8 pm were more likely to experience recurrent malaria episodes (aOR, 8.31,95%CI, 2.21-31.27 P=0.003).

**Table 4:** Shows the factors associated with recurrent malaria episodes among children under five at Kayunga Regional Referral Hospital.

| Variable | cOR | P value | aOR | P value |
| --- | --- | --- | --- | --- |
| **Period sprayed** |  |  |  |  |
| Monthly | Ref |  | Ref |  |
| Annually | 0.23(0.09-0.58) | **<0.001\*\*\*** | 8.93(2.11-10.81) | **0.003\*\*** |
| Others | 1.91(0.63-5.77) | 0.26 | 0.78(0.12-5.20) | 0.812 |
| I don't spray | 0.22(0.05-1.05) | 0.06 | 3.80(1.27-9.41) | 0.017 |
| **Anti-malaria commonly used to treat malaria.** |  |  |  |  |
| Coartem | Ref |  | Ref |  |
| Quinine | 0.86(0.18-4.04) | 0.85 | 0.28(0.12-0.65) | **0.003\*\*** |
| Artesunate | 0.23 (0.00-0.02) | **0.011\*** | 0.61(0.17-2.19) | 0.446 |
| p-alaxin | 0.63(0.10-3.82) | 0.61 | 2.22(0.39-8.65) | 0.366 |
| **At what time do you close windows every day** |  |  |  |  |
| 3-4 pm | Ref |  | Ref |  |
| 5-6 pm | 1.27(0.13-12.03) | 0.84 | 2.11(0.36-12.96) | 0.721 |
| 7-8 pm | 2.19(1.10-4.36) | **0.031\*** | 8.31(2.21-10.27) | **0.002\*\*** |
| *\*level of significance at $p<0.05$, \*\*\*level of significance at $p<0.001$, $p\leq 0.05$* |  |  |  |  |

## Discussion

Our results showed an 84% prevalence of recurrent malaria episodes in children under five, a notably high figure compared to other studies. For instance, Aongart and colleagues reported a 30% prevalence of malaria recurrences within 28 days after the initial treatment [22]. Similarly, a study conducted in Western Thailand found that 9.78% of individuals were infected with malaria two or more times in 2019 [23]. The lower prevalence of recurrent malaria in these studies can be attributed to adherence to preventive practices such as indoor residual spraying, avoiding self-medication (which can lead to antimalarial drug resistance), and preventing childhood illnesses like measles and chickenpox, which weaken children’s immunity and make them more vulnerable to malaria. These measures collectively reduce malaria recurrence risk by addressing environmental factors and individual susceptibility.

Our findings indicate that children from households that were annually sprayed were more likely to experience recurrent malaria episodes (aOR: 8.93, 95% CI: 2.11-10.81, p = 0.003). This contrasts with a study conducted in Northern Ghana, which found that indoor residual spraying reduced the chances of recurrent malaria infections in children under five [24]. The discrepancy between these studies could be attributed to differences in the study settings and other behaviours related to malaria prevention. According to Nana-Kwadwo Biritwum and colleagues (2019), children under five in Ghana were using both indoor residual spraying and insecticide-treated mosquito nets, which provided additional protection from mosquito bites and consequently reduced malaria incidence[24]. This suggests that indoor residual spraying for mosquito prevention should be complemented with other malaria prevention methods, particularly for children under the age of five, to be more effective.

Our results indicate that children treated with quinine in a previous malaria infection were less likely to experience recurrent malaria episodes than those who used other anti-malarial medications (aOR: 0.28, 95% CI: 0.12-0.65, p = 0.003). This finding is consistent with a study conducted in Indonesia, which showed that the use of quinine reduces the chances of malaria infection reoccurrence [25]. This efficacy is likely due to quinine’s continued effectiveness against drug-resistant Plasmodium falciparum strains, ensuring successful treatment [26]. These results underscore the importance of continuing to use quinine as a treatment for malaria, particularly in areas where drug resistance is prevalent.

Our results show that children who lived in houses with windows closed as late as 7 p.m. - 8 p.m. were more likely to have recurring malaria episodes compared to those who had windows closed earlier (aOR: 8.31, 95% CI: 2.21-31.27, p = 0.003). This finding shows the critical importance of the timing of window closure in malaria prevention. Similar studies have indicated that the practice of closing doors and windows is significantly associated with malaria transmission [27,28]. Anopheles mosquitoes are most active during dusk and early evening hours, so closing windows later in the evening exposes children to mosquito bites at a critical time, increasing the likelihood of malaria transmission [29,30]. This extended exposure period can result in more frequent malaria episodes. In Northern Uganda, especially during hot nights in the dry season, people might delay closing windows for ventilation, inadvertently increasing malaria risk [31]. This study highlights the need for community education on the importance of closing windows before dusk as a simple yet effective measure to prevent malaria.

## Conclusion

Nearly 90% of children under five experienced recurrent malaria. These infections were more common in homes with annual insecticide spraying and delayed window closures than in those treated with quinine-based antimalarials. While quinine is an important treatment, alternative prevention strategies like regular indoor spraying and timely window closure should be prioritized.

## Data Availability

Data cannot publicly be shared because of fear of data misuse or replication without giving proper credit. Contact the corresponding author via email at to get a copy of the data files

## Acknowledgements

We acknowledge the invaluable contribution of Lira University and our research participants to this study.

## REFERENCES

1. Ale S, Akande S, Organ R, Rauf Q. Mathematical Modeling for the Transmission Dynamics Control of HIV and Malaria Coinfection in Nigeria towards Attaining Millenium Development Goal. 2022;

2. WHO. Fact sheet about malaria [Internet]. 2023 [cited 2023 May 5]. Available from: https://www.who.int/news-room/fact-sheets/detail/malaria

3. WHO. World Malaria Report 2019 [Internet]. 2019 [cited 2024 Jun 14]. Available from: https://www.who.int/publications/i/item/9789241565721

4. Tsegaye AT, Ayele A, Birhanu S. Prevalence and associated factors of malaria in children under the age of five years in Wogera district, northwest Ethiopia: A cross-sectional study. PLOS ONE [Internet]. 2021 [cited 2023 Jun 18];16:e0257944. Available from: https://journals.plos.org/plosone/article?id=10.1371/journal.pone.0257944

5. WHO, 2019 [Internet]. [cited 2022 Jul 20]. Available from: https://www.who.int/news/item/23-04-2020-who-urges-countries-to-move-quickly-to-save-lives-from-malaria-in-sub-saharan-africa

6. Mpimbaza A. The age-specific incidence of hospitalized paediatric malaria in Uganda. BMC Infect Dis [Internet]. 2020 [cited 2023 Jun 27];20:503. Available from: 10.1186/s12879-020-05215-z

7. Kotepui M, Punsawad C, Kotepui KU, Somsak V, Phiwklam N, PhunPhuech B. Prevalence of malarial recurrence and haematological alteration following the initial drug regimen: a retrospective study in Western Thailand. BMC Public Health [Internet]. 2019 [cited 2023 Jun 18];19:1294. Available from: 10.1186/s12889-019-7624-1

8. Guyant P, Canavati SE, Chea N, Ly P, Whittaker MA, Roca-Feltrer A, et al. Malaria and the mobile and migrant population in Cambodia: a population movement framework to inform strategies for malaria control and elimination. Malar J [Internet]. 2015 [cited 2023 Jul 17];14:252. Available from: 10.1186/s12936-015-0773-5

9. Wilson AL, Bradley J, Kandeh B, Salami K, D’Alessandro U, Pinder M, et al. Is chronic malnutrition associated with an increase in malaria incidence? A cohort study in children aged under 5 years in rural Gambia. Parasit Vectors [Internet]. 2018 [cited 2023 Jul 17];11:451. Available from: 10.1186/s13071-018-3026-y

10. Foolchand A, Ghazi T, Chuturgoon AA. Malnutrition and Dietary Habits Alter the Immune System Which May Consequently Influence SARS-CoV-2 Virulence: A Review. Int J Mol Sci [Internet]. 2022 [cited 2024 Jun 14];23:2654. Available from: https://www.ncbi.nlm.nih.gov/pmc/articles/PMC8910702/

11. Bresnahan KA, Tanumihardjo SA. Undernutrition, the Acute Phase Response to Infection, and Its Effects on Micronutrient Status Indicators. Adv Nutr [Internet]. 2014 [cited 2023 Jul 17];5:702–11. Available from: 10.3945/an.114.006361

12. MOH. National Malaria Control Program [Internet]. Minist. Health Gov. Uganda. 2020 [cited 2024 Jun 11]. Available from: https://www.health.go.ug/programs/national-malaria-control-program/

13. Kamya MR, Arinaitwe E, Wanzira H, Katureebe A, Barusya C, Kigozi SP, et al. Malaria Transmission, Infection, and Disease at Three Sites with Varied Transmission Intensity in Uganda: Implications for Malaria Control. Am J Trop Med Hyg [Internet]. 2015 [cited 2023 Jun 19];92:903–12. Available from: https://www.ncbi.nlm.nih.gov/pmc/articles/PMC4426576/

14. Prevention C-C for DC and. CDC - Malaria - About Malaria - Disease [Internet]. 2022 [cited 2023 Jun 16]. Available from: https://www.cdc.gov/malaria/about/disease.html

15. MOH. Malaria indicator survey 2018-19 [Internet]. 2020 [cited 2023 Jun 19]. Available from: https://www.google.com/search?q=According+to+the+2020+Uganda+Malaria+Indicator+Survey&oq=According+to+the+2020+Uganda+Malaria+Indicator+Survey&aqs=chrome..69i57j33i160l3.1255j0j7&sourceid=chrome&ie=UTF-8

16. Westercamp N, Staedke SG, Maiteki-Sebuguzi C, Ndyabakira A, Okiring JM, Kigozi SP, et al. Effectiveness of in-service training plus the collaborative improvement strategy on the quality of routine malaria surveillance data: results of a pilot study in Kayunga District, Uganda. Malar J [Internet]. 2021 [cited 2024 Jul 19];20:290. Available from: https://malariajournal.biomedcentral.com/articles/10.1186/s12936-021-03822-y

17. Zalwango MG, Bulage L, Zalwango JF, Migisha R, Agaba BB, Kadobera D, et al. Trends and Distribution of Severe Malaria Cases, Uganda, 2017–2021: Analysis of Health Management Information System Data. [cited 2024 Jul 19]; Available from: https://uniph.go.ug/wp-content/uploads/2023/07/Trends-and-Distribution-of-Severe-Malaria-Cases-Uganda-2017%E2%80%932021-Analysis-of-Health-Management-Information-System-Data.pdf

18. Nalikka O. Case management practices for children under five with severe malaria: facilitators and barriers to appropriate care at Kayunga Hospital [Internet] [PhD Thesis]. Makerere university; 2022 [cited 2024 Jul 19]. Available from: http://makir.mak.ac.ug/handle/10570/10157

19. kish and Leslie formula [Internet]. 1995 [cited 2023 Jun 19]. Available from: https://www.google.com/search?sxsrf=APwXEdf_tA-j0klKuLAARt8rNciUccmmjA:1687194464786&q=Leslie+Kish+formula+FOR+DETERMINING+SAMPLE+SIZE&tbm=isch&sa=X&ved=2ahUKEwiq_vWV6c__AhWxVqQEHc3ZC8IQ0pQJegQICxAB&biw=1366&bih=657&dpr=1

20. Simões LR, Alves Jr ER, Ribatski-Silva D, Gomes LT, Nery AF, Fontes CJF. Factors associated with recurrent plasmodium vivax malaria in Porto Velho, Rondônia state, Brazil, 2009. Cad Saude Publica [Internet]. 2014 [cited 2024 Jun 8];30:1403–17. Available from: https://www.scielo.br/j/csp/a/7ktTQK4C834xVdwxTTRtPhm/

21. Lawpoolsri S, Sattabongkot J, Sirichaisinthop J, Cui L, Kiattibutr K, Rachaphaew N, et al. Epidemiological profiles of recurrent malaria episodes in an endemic area along the Thailand-Myanmar border: a prospective cohort study. Malar J [Internet]. 2019 [cited 2023 Jul 17];18:124. Available from: 10.1186/s12936-019-2763-5

22. Mahittikorn A, Masangkay FR, Kotepui KU, Milanez GDJ, Kotepui M. The high risk of malarial recurrence in patients with Plasmodium-mixed infection after treatment with antimalarial drugs: a systematic review and meta-analysis. Parasit Vectors [Internet]. 2021 [cited 2023 Jun 28];14:280. Available from: 10.1186/s13071-021-04792-5

23. Kotepui M, Punsawad C, Kotepui KU, Somsak V, Phiwklam N, PhunPhuech B. Prevalence of malarial recurrence and haematological alteration following the initial drug regimen: a retrospective study in Western Thailand. BMC Public Health [Internet]. 2019 [cited 2024 Apr 24];19:1294. Available from: 10.1186/s12889-019-7624-1

24. Nana-Kwadwo Biritwum, et al. Impact of Indoor Residual Spraying on Malaria Incidence and Prevalence in Children Less Than Five Years of Age in Northern Ghana. Trop Parasitol [Internet]. 2019 [cited 2024 Apr 24];6:30–41. Available from: https://www.ncbi.nlm.nih.gov/pmc/articles/PMC4778180/

25. Sikora SA, Poespoprodjo JR, Kenangalem E, Lampah DA, Sugiarto P, Laksono IS, et al. Intravenous artesunate plus oral dihydroartemisinin–piperaquine or intravenous quinine plus oral quinine for optimum treatment of severe malaria: lesson learnt from a field hospital in Timika, Papua, Indonesia. Malar J [Internet]. 2019 [cited 2024 Jun 8];18:1–12. Available from: https://link.springer.com/article/10.1186/s12936-019-3085-3

26. Aminake MN, Pradel G. Antimalarial drugs resistance in Plasmodium falciparum and the current strategies to overcome them. Microb Pathog Strateg Combat Them Sci Technol Educ [Internet]. 2013 [cited 2024 Jun 8];1:269–82. Available from: https://www.researchgate.net/profile/Nigel-Makoah/publication/280717864_Antimalarial_drugs_resistance_in_Plasmodium_falciparum_and_the_current_strategies_to_overcome_them/links/55c248d508aeb975673e4012/Antimalarial-drugs-resistance-in-Plasmodium-falciparum-and-the-current-strategies-to-overcome-them.pdf

27. Ngadjeu CS, Doumbe-Belisse P, Talipouo A, Djamouko-Djonkam L, Awono-Ambene P, Kekeunou S, et al. Influence of house characteristics on mosquito distribution and malaria transmission in the city of Yaoundé, Cameroon. Malar J [Internet]. 2020 [cited 2024 Jun 8];19:53. Available from: 10.1186/s12936-020-3133-z

28. Mburu MM, Juurlink M, Spitzen J, Moraga P, Hiscox A, Mzilahowa T, et al. Impact of partially and fully closed eaves on house entry rates by mosquitoes. Parasit Vectors [Internet]. 2018 [cited 2024 Jun 8];11:383. Available from: 10.1186/s13071-018-2977-3

29. Mwema T. Anopheles vectors species composition, their biting cycle and role of human behaviour in Malaria transmission in an endemic region from Namibia [Internet] [PhD Thesis]. University of Namibia; 2021 [cited 2024 Jul 19]. Available from: https://repository.unam.edu.na/handle/11070/3000

30. Takken W, Charlwood D, Lindsay SW. The behaviour of adult Anopheles gambiae, sub-Saharan Africa’s principal malaria vector, and its relevance to malaria control: a review. Malar J [Internet]. 2024 [cited 2024 Jul 19];23:161. Available from: https://malariajournal.biomedcentral.com/articles/10.1186/s12936-024-04982-3

31. Ababa A. National malaria guidelines. 2018 [cited 2024 Jul 19]; Available from: https://doctorsonlinee.com/wp-content/uploads/2022/04/National_malaria_guideline_2018-12-1-1.pdf

